# Understanding the factors affecting global political priority for controlling sexually transmitted infections: a qualitative policy analysis

**DOI:** 10.1101/2023.10.03.23296476

**Authors:** Dadong Wu, Nicola Low, Sarah J Hawkes

**Affiliations:** Affiliated Shenzhen Maternity & Child Healthcare Hospital, Southern Medical University, Shenzhen, China; Center for World Health Organization Studies, School of Health Management of Southern Medical University, Guangzhou, China; Institute of Social and Preventive Medicine (ISPM), University of Bern, Bern, Switzerland; Institute for Global Health, University College London, London, UK

**Keywords:** sexually transmitted infections, global health agenda, political priority, informant interview, policy analysis

## Abstract

**Introduction:** Sexually transmitted infections (STIs) are a significant public health challenge, but there is a perceived lack of political priority in addressing STIs as a global health issue. Our study aimed to understand the determinants of global political priority for STIs since the 1980s and to discern implications for future prioritisation.

**Methods:** Through semi-structured interviews from July 2021 to February 2022, we engaged 20 key stakeholders (8 women, 12 men) from academia, United Nations agencies, international non-governmental organisations, philanthropic organisations, and national public health agencies. A published policy framework was employed for thematic analysis, and findings triangulated with relevant literature and policy documents. We examined issue characteristics, prevailing ideas, actor power dynamics and political contexts.

**Results:** A contrast in perspectives before and after the year 2000 emerged. STI control was high on the global health agenda during the late 1980s and 1990s, as a means to control HIV. A strong policy community agreed on evidence about the high burden of STIs and that STI management could reduce the incidence of HIV. The level of importance decreased when further research evidence did not find an impact of STI control interventions on HIV incidence. Since 2000, cohesion in the STI community has decreased. New framing for broad STI control has not emerged. Interventions that have been funded, such as human papillomavirus vaccination and congenital syphilis elimination have been framed as cancer control or improving newborn survival, rather than as STI control.

**Conclusion:** Globally, the perceived decline in STI control priority might stem from discrepancies between investment choices and experts’ views on STI priorities. Addressing STIs requires understanding the intertwined nature of politics and empirical evidence in resource allocation. The ascent of universal health coverage presents an opportunity for integrated STI strategies but high-quality care sustainable funding and strategic coordination are essential.

**Key messages:** *What is already know on this topic?:*

■ Setting priorities within health services is a political process.
■ Sexually transmitted infections (STIs) other than HIV are a significant public health issue.
■ STI control was high on the global health agenda in the late 1980s and 1990s, when it was promoted as a means to lower the transmission of HIV, but attention paid to STI control appears to have waned over the past two decades.

*What this study adds?:*

■ A range of factors, including but not limited to, empirical evidence of disease burden, have driven the attention paid to STI control over time.
■ The STI community has lacked cohesion, champions and engagement with civil society, thus contributing to their lowered position on health policy agendas.
■ STI control has been successful when framed as reaching aligned goals in other areas – HIV control, maternal and child health, cancer control – or when a biomedical intervention (vaccine, diagnostic) is available.

*How this study might affect research, practice or policy?:*

■ A more politically aware approach to STI control could increase policy attention and resource allocation, moving beyond technical evidence to understanding and leveraging political context.
■ The rollout of universal health coverage can present opportunities to integrate STI control into broader health policy reform and prioritisation, but the STI community will need to pay attention to issue-framing, community cohesion, and the role of policy entrepreneurs if they are to have success in forging a window of policy opportunity.
■ STI advocacy needs to be strengthened through strategic alliances with a diverse group of stakeholders, including civil society (e.g., those representing the broader sexual and reproductive health agenda and the cancer agenda).

## Introduction

Setting priorities within health services is a political process – driven not just by evidence of the burden of any particular condition, but also by the power of policy actors, prevailing ideas, and the emergence of windows of opportunity.^1,2^ At the global level, political priority refers to “the degree to which international and national political leaders actively give attention to an issue and back up that attention with financial, technical, and human resources that are commensurate with the issue’s severity.”^3^ The relative position of any health issue on the global health agenda also reflects the importance of social values and issue-framing, which drive the attention paid to the issue.^4^ There is a perception that global attention to the control of sexually transmitted infections (STIs) other than HIV is insufficient^5^ and has declined since the late 1980s and 1990s,^6^ when STI control was promoted as a means to lower the transmission of HIV.^7–9^ The term STIs comprises a range of infections, many of which are common and, together, cause substantial morbidity and mortality. The World Health Organization (WHO) estimates that there were 374 million new cases of four curable infections (chlamydia, gonorrhoea, syphilis, and trichomoniasis) in 2020^10^. According to the 2019 Global Burden of Disease (GBD) study, non-HIV STIs were associated with 8.57 million disability-adjusted life years (DALYs), of which 62.3% can be attributed to neonatal syphilis.^11^ The GBD estimates of STI burden would increase if conditions such as human papillomavirus (HPV) infection, which causes most cervical cancer^12^and the contribution of STIs to conditions such as preterm birth, were included.

The question of whether and why STI control really has dropped down the policy agenda has not been examined systematically but is of interest and importance for those seeking to ensure appropriate and fair levels of resource allocation to achieve goals of STI control because resources are limited. Ideally, this task should be a collaborative effort, shared between “the Ministry of Health and the entire health stakeholder community” including citizens and health system providers. One indicator of relative priority is financial resource allocation.^13^ Grollman et al. reported that the four curable STIs accounted for 16% (US$ 693 million) of total official development assistance (ODA) and grants from the Bill & Melinda Gates Foundation allocated to reproductive, maternal, newborn, and child health in 2003. However, this percentage declined to 1% by 2006 and remained at this level, amounting to US$ 83 by 2013.^14^ WHO estimated a need of US$ 18,200 million for global STI prevention and control efforts in over 100 low– and middle-income countries (LMICs) between 2016 and 2021.^15^ It is not yet clear what proportion of this amount was allocated, but there are thought to be significant funding gaps, from both ODA allocations and contributions at the national ministry level in many settings.^16^ Some specific interventions have gained priority on the global health agenda. For example, the Global Fund to Fight AIDS, Tuberculosis and Malaria invested US$3.12 billion between 2003 and 2010 in maternal, newborn and child health, which includes prevention of mother-to-child of transmission (PMTCT) of syphilis.^17^ Gavi, the Vaccine Alliance committed up to US$ 500 million to support the introduction of HPV vaccination in 40 LMICs from 2016-2020.^18^ Also, the Global Antibiotic Research and Development Partnership invested €75 million in 2021 into developing new treatments for antimicrobial-resistant infections, including gonorrhoea.^19^

In this paper, we seek to understand the determinants of global political priority for STIs over the past four decades (1980-2022) and to discuss the implications for future priority setting.

## Methods

### Study design

To undertake this qualitative policy analysis, we triangulated evidence from interviews with key informants and from a review of published studies, organisational reports and grey literature. We report our findings according to the Consolidated Criteria for Reporting Qualitative Research^20^ and Sex and Gender Equity in Research guidelines.^21^

### Policy framework

Analysis and synthesis of qualitative data were guided by a conceptual framework developed by Shiffman and Smith to determine global political priority of health issues.^3^ The framework comprises four categories, which cover eleven determinants of political priority (table 1) and has been applied to the analysis of a number of global health initiatives, such as maternal mortality reduction, mental health, global surgery, emergency care, and early childhood development.^3,22–25^

**Table 1.** The four categories of determinants of global political priority.

| Category | Description |
| --- | --- |
| Issue characteristics | Features of the problem |
| Ideas | The ways in which those involved with the issues understand and portray it |
| Actor power | The strength of the individuals and organisations concerned with the issue |
| Political contexts | The environments in which actors operate |

From Shiffman and Smith<sup>3</sup>

The category of the issue characteristics category looks at the nature of the issue itself. Problems that can be measured by credible indicators are more likely to attract attention as policymakers and funders will have information to confirm the severity and monitor progress.^26^ Moreover, policymakers are more inclined to address a problem if there are effective interventions.^27^ The category of ideas examines how an issue and its solution are understood and portrayed both within the policy community and publicly – the frame.^28^ Actor power considers the performance of networks comprising individuals from various organisations who share a common policy concern. The membership, structure and organisation of these policy communities determine their impact on the policy processes.^27,29^ Global and national policy communities function more effectively in shaping policy agendas where influential entrepreneurs or strong guiding institutions emerged to lead them.^27,30^ Additionally, initiatives that connect with grassroots organisations in civil society are more likely to obtain policy attention.^3^ Finally, the category of political contexts explores the environment in which actors operate, especially “policy windows” which refer to the key moments when conditions align favourably for certain issues, as well as global governance structure in the sector.^27^

### Data collection

#### Informant interviews

We conducted a stakeholder mapping^31^ in June 2021 to guide identification of potential informants, based on our experiences in STI-related research and from publications. We also followed informants’ referrals across multiple domains, including funders, policy makers, advocates and researchers. We aimed for gender balance in selecting interview respondents. Potential informants were contacted using a standardised email, which explained the purpose of the study, potential risks, and how privacy and confidentiality would be maintained. All respondents signed a consent form allowing audio recording of interviews and had the opportunity to ask questions before the start of the interview (online supplemental file 1a, 1b).

Semi-structured interviews were used, following a general interview guide based on the Shiffman and Smith framework. Owing to COVID-19 international travel restrictions, in-person interviews were not feasible. DW, an early-career female researcher, with experience in health policy analysis and STI control, conducted all discussions in English via online platforms. The researcher had no prior personal relationships with the informants. Each interview involved only the interviewer and participant and lasted 30 to 90 minutes, during which notes were taken. No repeat interviews were conducted. Questions were tailored for each informant based on their position and responsibilities around STI control. If feasible, they were also invited to comment on anonymised answers of other respondents. To assess power dynamics and their evolution over time, informants were asked to identify key actors shaping the global health agenda and influencing resource allocation. At the end of each interview, they were queried on the most influential factor for prioritisation of STIs. Respondent recruitment persisted until theoretical saturation was achieved, i.e. when all factor themes had been identified and additional interviews were unlikely to reveal new information.^32^

The recorded interviews were transcribed and all materials were stored digitally in password-protected computers and de-identified during data analysis. Transcripts were not sent back to informants, but some were contacted to ensure the accuracy of quotes.

#### Literature review

We performed a literature review concurrently with the interviews. We collected data about global policies and practices for STI control by searching established databases and websites of organisations involved in advocating for and/or financing STI control. We searched PubMed, Web of Science, and Google Scholar to identify relevant studies published in English between January 1980 and December 2022. The search strings combined MeSH headings 2022 (“sexually transmitted diseases”, “syphilis”, “gonorrhea”, “chlamydia infections”, “trichomonas”, “herpes genitalis”, “human papillomavirus”) and free-text terms (“policy”, “priority”, “salience”, “prioritisation”, “agenda setting”, “decision making”, “policy making”). We also searched the WHO Library and websites of three United Nations agencies (Joint United Nations Programme on HIV/AIDS, United Nations Children’s Fund, United Nations Population Fund), the Global Fund for AIDS, TB and Malaria, and the Bill and Melinda Gates Foundation. In addition, some informants directed us to particular projects and studies. We selected articles and documents based on their relevance to the political prioritisation of STI control.

### Data analysis and synthesis

Using the four categories and eleven factors from the Shiffman and Smith framework as main themes and sub-themes, we conducted an iterative thematic analysis.^33^ The NVivo software (version 11) was employed to organise and analyse the interview transcripts. A single researcher (DW) coded all the transcripts, and the findings were cross verified with one another as well as against published studies and organisational reports. When reporting the interview findings, we assigned each key informant a number and cited relevant literature and documents from our review to give a broader interpretation and contextualisation of the interview findings. During the analysis, the findings were discussed via online meetings with other researchers (NL and SH) and at a face-to-face meeting in June 2022 involving the multidisciplinary project team (online supplemental file 2).

### Patient and Public Involvement

This study was part of a multidisciplinary project examining the political prioritisation of the prevention and control of STIs (online supplemental file 2).^34^ No patients participated in the design or conduct of this policy analysis. As part of the larger project, we did interview pregnant –18 and healthcare workers in Papua New Guinea and Zambia to explore civil society mobilisation and advocacy and we report on their priorities, experiences, or preferences separately.

**Ethics approval** The Cantonal Research Ethics Committee in Bern, Switzerland (Req-2020-00269, March 2020) determined that the study was exempt from the Human Research Act, Art. 2, Paragraphs 1, Switzerland (online supplemental file 3).

## Results

From July 2021 to February 2022, we contacted 34 potential informants, of whom 23 responded and 3 declined to be interviewed (59% acceptance rate). Of the 20 respondents, 8 were women, only 2 were originally from LMICs and 15 first became involved in STI control and prevention before 2000 (Table 2). The respondents came from 10 countries (US, Zimbabwe, Belgium, Netherland, UK, Bangladesh, Australia, France, Italy, Switzerland) and have worked in different types of organisations, including United Nations agencies (WHO headquarters or regional offices, Joint United Nations Programme on HIV/AIDS), national public health agencies, development partners (bilateral assistance programmes, private philanthropic funders), international non-governmental organisations (NGOs), and academia.

**Table 2.** Characteristics of key informants.

| Informant | Gender | First involvement in STI control | Type of primary affiliation | Income category of country of origin |
| --- | --- | --- | --- | --- |
| 1 | Man | Before 2000 | Academia | High |
| 2 | Man | Before 2000 | United Nations agency | Low |
| 3 | Woman | Before 2000 | Academia | High |
| 4 | Man | Before 2000 | Bilateral assistance programme | High |
| 5 | Woman | Before 2000 | Academia | High |
| 6 | Man | Before 2000 | Academia | High |
| 7 | Woman | Before 2000 | Philanthropic funder | High |
| 8 | Man | Before 2000 | United Nations agency | Low |
| 9 | Man | Before 2000 | International NGO | High |
| 10 | Man | After 2000 | Philanthropic funder | High |
| 11 | Man | Before 2000 | United Nations agency | High |
| 12 | Man | After 2000 | International NGO | High |
| 13 | Man | Before 2000 | Academia | High |
| 14 | Woman | Before 2000 | Academia | High |
| 15 | Man | Before 2000 | Academia | High |
| 16 | Woman | Before 2000 | Academia | High |
| 17 | Man | After 2000 | International NGO | High |
| 18 | Woman | Before 2000 | Academia | High |
| 19 | Woman | After 2000 | National public health<br>agency | High |
| 20 | Woman | After 2000 | United Nations agency | High |

NGO: non-governmental organisation

We report our findings about factors affecting actor power, ideas, political contexts and issue characteristics, particularly contrasting the periods before and since 2000. This timeframe emerged from the interview data as the approximate timing of an apparent shift in donor attention on STI control.

### Issue characteristics

#### Before 2000

The World Bank’s 1993 World Development Report stated that STIs, excluding HIV, accounted for 9% of the disease burden among adult women and 2% among adult men.^7^ This report emphasised the cost-effectiveness of treating bacterial STIs, playing a crucial role in raising awareness about the burden of STIs and the importance for addressing their control. This contributed to STIs being portrayed as a “tremendous public health problem” deserving policy, donor, and research attention in the early 1990s (I4, I8). Syndromic management to treat the most common causes of STI symptoms gained ground at the primary care level in Africa, where there were no simple and accurate diagnostic tests for most STIs (I5, I6, I7, I8, I13 and I14).^35^ As a respondent stated:

> *“…pointed to the necessity of non-specialist approaches so much more decentralised approaches to STI diagnosis and management and, in doing so, help to raise the profile if you wish of this public health problem.” (I4)*

#### Since 2000

Estimates of global STI burden have been contested by some informants (I2, I11, I18 and I20) for two reasons: First, the underlying basis of the estimates is uncertain because data such as prevalence, incidence, mortality, and antimicrobial resistance patterns of STIs remain unknown in many settings with poor information collection and surveillance.^36,37^ Second, many burden assessments have not included all STI-associated impacts such as HPV-related cancers and neonatal morbidity and mortality. Informants attributed the persisting “unclear magnitude” of STIs worldwide to a chronic lack of funding for epidemiological research (I2, I4, I7, I8, I16 and I20). As one mentioned:

> *“[T]o some extent, you have these…self-reinforcing systems or vicious circles where the lack of funding results in a lack of data and a lack of data makes everybody think that there is no problem, and that leads to even less funding.” (I4)*

Evidence from the late 1990s raised concerns about the effectiveness and cost-effectiveness of syndromic management as a means to treat STIs, further decreasing the policy options available for STI control.^38,39^ Two respondents highlighted the lack of clear interventions (I11 and I19), noting that WHO set “aspirational” targets, such as reducing syphilis and gonorrhoea incidence by 90% by 2030, without providing countries with specific guidance on how to achieve these goals. Many informants attributed the neglect of STIs to the dearth of affordable diagnostics and treatments in LMICs (I5, I7, I11, I13, I14 and I18), leaving syndromic management as the main intervention for STI control, despite its problems. Congenital syphilis control is an exception because there is robust evidence of the effectiveness and cost-effectiveness of screening in pregnancy and scaling up is being facilitated by innovative tools, such as dual rapid tests for HIV and syphilis.

> *“There’s a renewed interest in STD control, however, we are still stuck with the absence of point-of-care testing…So the problem has not gone away and certainly not been solved.” (I13)*

### Ideas and issue framing

#### Before 2000

In the 1980s and early 1990s as evidence of the substantial impact of HIV on people and economies became clear, there was an active hunt for affordable and effective solutions to the HIV crisis. Epidemiological synergy between STIs and HIV was revealed^8^ and randomised controlled trials to examine the effect of interventions to control STIs on HIV transmission were launched.^40^ The first trial, published in 1995, found that communities provided with STI syndromic management in Mwanza Region, Tanzania had a lower incidence of HIV infection than communities without STI control (the trial is widely referred to as “the Mwanza trial”).^9^

Many respondents agreed that the Mwanza trial findings greatly enhanced the prioritisation of STIs (I2, I3, I4, I5, I6, I7, I8 and I9). Syndromic management was then portrayed as a means of tackling the HIV epidemic, including as part of an integrated reproductive health programme^41^ reaching women in family planning and antenatal clinics.^42^

> *“It was even believed at a certain point that STI control was the magic bullet for HIV prevention.” (I3)*

However, another randomised controlled trial, published in 1998, found no impact on HIV transmission of mass antimicrobial treatment for STIs at the village level in Rakai, Uganda (referred to as “the Rakai trial”).^43,44^ Additionally, two informants noted that the advent of highly active antiretroviral therapy at the Vancouver AIDS conference in 1996 further reduced the relevance of other STIs in HIV control among global actors(I3 and I16).^45^ This led to growing scepticism about prioritising STI management as part of HIV control, especially among major donors (I1, I2, I3, I4, I5, I7, I9 and I19). Consequently, consensus among global actors diminished,^46^ prompting major donors to withdraw resources from STI management initiatives (I4, I5 and I9).

> *“PEPFAR [the United States President’s Emergency Plan for AIDS Relief] did not put money into it anymore. PEPFAR put all its money into HIV prevention, into antiretroviral treatment, male circumcision, and prevention of mother-to-child transmission.” (I5)*

#### Since 2000

The findings of the Rakai trial in 1998, along with other trials of syndromic management and suppression of herpes simplex published since 2000,^47–49^ changed the balance of scientific opinion about the linkage between STI management and HIV control. This shift was compounded by the STI community’s failure to establish alternative framings that were powerful enough to sustain policy attention. As commented:

> *“The linkage to HIV was our biggest chance to have an integrated approach to control all the STIs and HIV. And I think we placed too much emphasis on that so that when the data didn’t support this as a co-intervention for prevention of HIV that there was a loss of interest.” (I19)*

The informants shared the view that prioritisation of non-HIV STIs since 2000 has been hindered by the popular perception that they are treatable, not fatal, and have a significantly lower burden than other major infectious diseases, like HIV and tuberculosis (I5, I8, I13, I14, I18 and I20), as well as the associated stigma of infections transmitted through sexual activity (I1, I4, I8, I9 and I20). Congenital syphilis was identified as an exception (I17, I19 and I20), with its framing as a major cause of stillbirths and neonatal deaths successfully stimulating international policy and donor attention.

Despite the widespread consensus within the policy community that some STIs are seriously neglected, there were significant differences in opinion on how to make a good investment case. Potential strategies suggested by the informants include: framing STIs as disproportionately affecting women’s health in the context of the MeToo movement against sexual abuse and harassment, which has “a different threshold for thinking about gender equity” (I7 and I16); framing STIs towards sexual and reproductive health and rights for all to “destigmatise STIs and take them out of a special realm” (I1, I2, I3, I5); and framing STIs as affecting key populations, especially those eligible for pre-exposure prophylaxis (PrEP) to prevent HIV infection (I1, I5, I7 and I9). No single issue emerged as a dominant framing for prioritising STI control during the interviews.

### Actor power

#### Before 2000

The number of policy actors working in STI control and prevention started to increase in the 1980s owing to rising concerns about studies in Africa that showed the high prevalence of bacterial STIs (I4, I8). WHO established a Venereal Diseases and Treponematoses Unit in 1986 and the Global Programme on AIDS (GPA) in 1987, both of which, according to Lush et al., hosted high profile meetings,^42^ culminating in a consensus statement with recommendations for coordinating AIDS and STI programmes.^50^

Two informants (I2 and I6) pointed to the strong leadership of GPA in promoting STI control in the 1990s, which helped secure support from key funders, such as the United States President’s Emergency Plan for AIDS Relief (PEPFAR), United States Agency for International Development, Department for International Development, UK, the World Bank, and others.^42^ The GPA became the Joint United Nations Programme on HIV/AIDS (UNAIDS) in 1996 and the funds were used to assist national governments in most African countries to introduce syndromic management guidelines through HIV control programmes (I2)^42^ and maternal and child health or family planning programmes.^38,41^ This process was facilitated by the active involvement of several international NGOs, such as Family Health International and the Population Council (I7 and I9). As one respondent put it:

> *“…HIV and STI colleagues at WHO would be amongst the most influential in terms of international policy in this area at that time.” (I6)*

The successful advocacy during this period was also attributed to the emergence of issue champions both in Africa and Europe. A key researcher and teacher at the Institute for Tropical Medicine Antwerp was identified by several respondents (I3, I4, I5 and I8) as playing a crucial role in the global policy community. Coined the “Antwerp Mafia”, the individual and many students and colleagues became influential in the STI and later HIV/AIDS communities, including in international organisations, such as WHO, UNAIDS, and the European Commission. These champions held strong authority and legitimacy due to their field experience and contacts and were able to allocate funds to STIs.^42^

#### Since 2000

In 1999, the STI Unit moved back from UNAIDS to WHO, joining the newly formed Division of Reproductive Health and Research (RHR), which signalled separation between the global STI and AIDS communities.^42^ Many respondents (I1, I5, I7, I10, I17 and I20) indicated that, since 2000, the STI community has been characterised by a loose structure and lack of champions. Although a group of policymakers, researchers, and programme managers worked closely with WHO’s RHR, forming a club-like camaraderie to develop STI control guidelines and strategies (I2),^51^ this group was mainly research-based and had limited impact on implementation at country level (I20). Two informants believed that the withdrawal of major donors had caused a so-called “brain drain” (I5 and I9), resulting in fewer young people with an interest in advocacy for STI control (I3 and I19), and personnel instability (I1 and I19) within the policy community. This has made the community less influential on the global health agenda.

> *“…some of the best people working in STI switched to HIV…these leaders were not just scientists, but also advocates who were very vocal…I think [that] has not helped for the STI world.” (I5)*

Furthermore, some informants (I15, I16 and I19) perceived that, due to scarce resources, WHO’s influence could hardly go beyond the creation of technical guidelines, thus diminishing its power in shaping the priority of STI control. This situation was accentuated by the lack of new effective coordinating mechanisms, especially when contrasted with the cohesive leadership of GPA in the 1990s.

During this period, the global STI control initiative has also been marked by weak mobilisation of civil society, with some informants citing insufficient funding as a reason (I17 and I20). Only two international NGOs, the Clinton Health Access Initiative and Evidence Action, were identified during the interviews as collaborating with WHO to support some African countries in implementing PMTCT of syphilis by providing technical assistance and fixing supply chain disruptions (I12, I17, I19 and I20).

> *“…what I’d highlight is having NGO partners that…have the capacity to support because…any time you’re sort of introducing a new service or refocusing priorities that it just requires a lot of change management and technical support.” (I12)*

Yet, even with the efforts of these NGOs, their reach and influence remained relatively limited in comparison to larger global health initiatives, like HIV/AIDS control.

### Political contexts

#### Before 2000

Given the importance of HIV and its framing as a health security threat which threatened economic and demographic stability in many parts of the world during the pre-2000 period,^52^ the initial evidence that STI control provided a solution for limiting HIV transmission provided an important policy window in the view of several respondents (I2, I3, I4, I5, I6 and I7).^42^ This window was effectively closed, with a consequent loss of attention and resources, when STI control was shown not to be effective at controlling HIV transmission.

#### Since 2000

Informants did not identify specific policy windows for the broad goal of STI control since 2000. However, published studies indicated that the global goals setting in the Millenium Development Goals (MDGs) in 2000 provided an opportunity to push for a focus on individual issues such as preventing congenital syphilis,^2,53^ with attendant impacts on MDG 4 (reducing child mortality), 5 (improving maternal health), and 6 (combating HIV/AIDS, malaria, and other diseases).^54^ Although advocates have successfully pushed for elements of STI control, such as PMTCT of syphilis, HPV vaccination, and treatment for drug resistant gonorrhoea, no specific global governance mechanism for STI control was identified during the interviews. Informants did not perceive the attention on specific interventions to be able to stimulate a broader focus or prioritisation of other STIs (I19 and I20). Meanwhile, although WHO has produced a number of technical global strategies for STI control since 2000,^15,55^ implementation was judged to be more likely in countries with robust governance capacity and adequate funding.^36,51^

The Sustainable Development Goals (SDGs) while not specifically mentioning STIs do provide opportunities to promote STI control in both SDG 5 (“universal access to comprehensive sexual and reproductive health and reproductive rights”) and SDG 3, (“ensuring healthy lives and promoting well-being for all”). The 2019, the United Nations General Assembly’s adoption of a new political declaration on universal health coverage (UHC), including commitments to increase investments in comprehensive sexual and reproductive healthcare services^56^ may open a policy window for STIs. According to an official from WHO:

> *“[What] we need to do with STIs is to better integrate it into primary care and UHC…because primary care is getting some funding. And therefore, we want STIs to be seen as an essential part of primary care.” (I20)*

Table 3 summarises the main factors that informants mentioned as affecting the global political priority of STIs in the Shiffman and Smith framework.

**Table 3.**
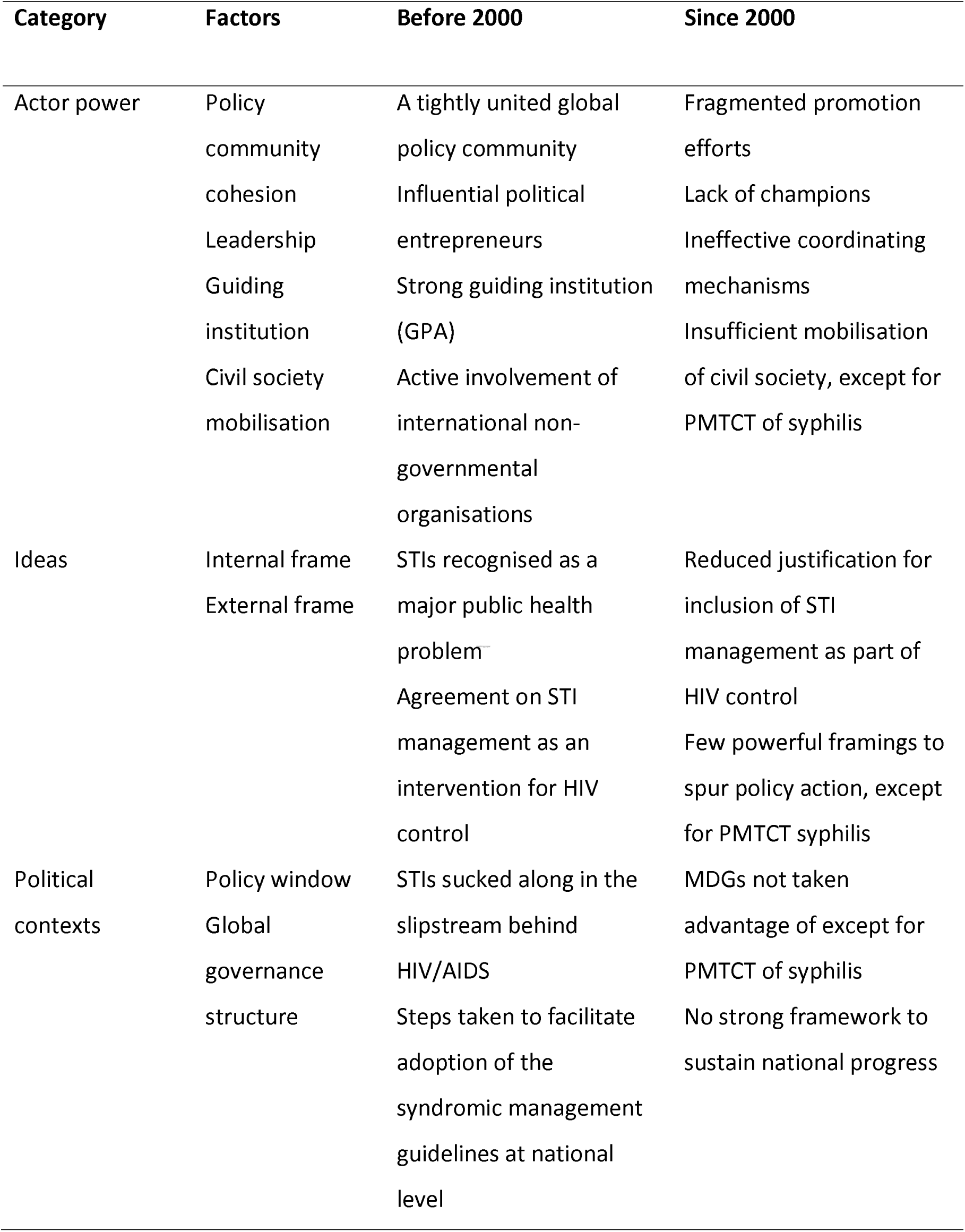
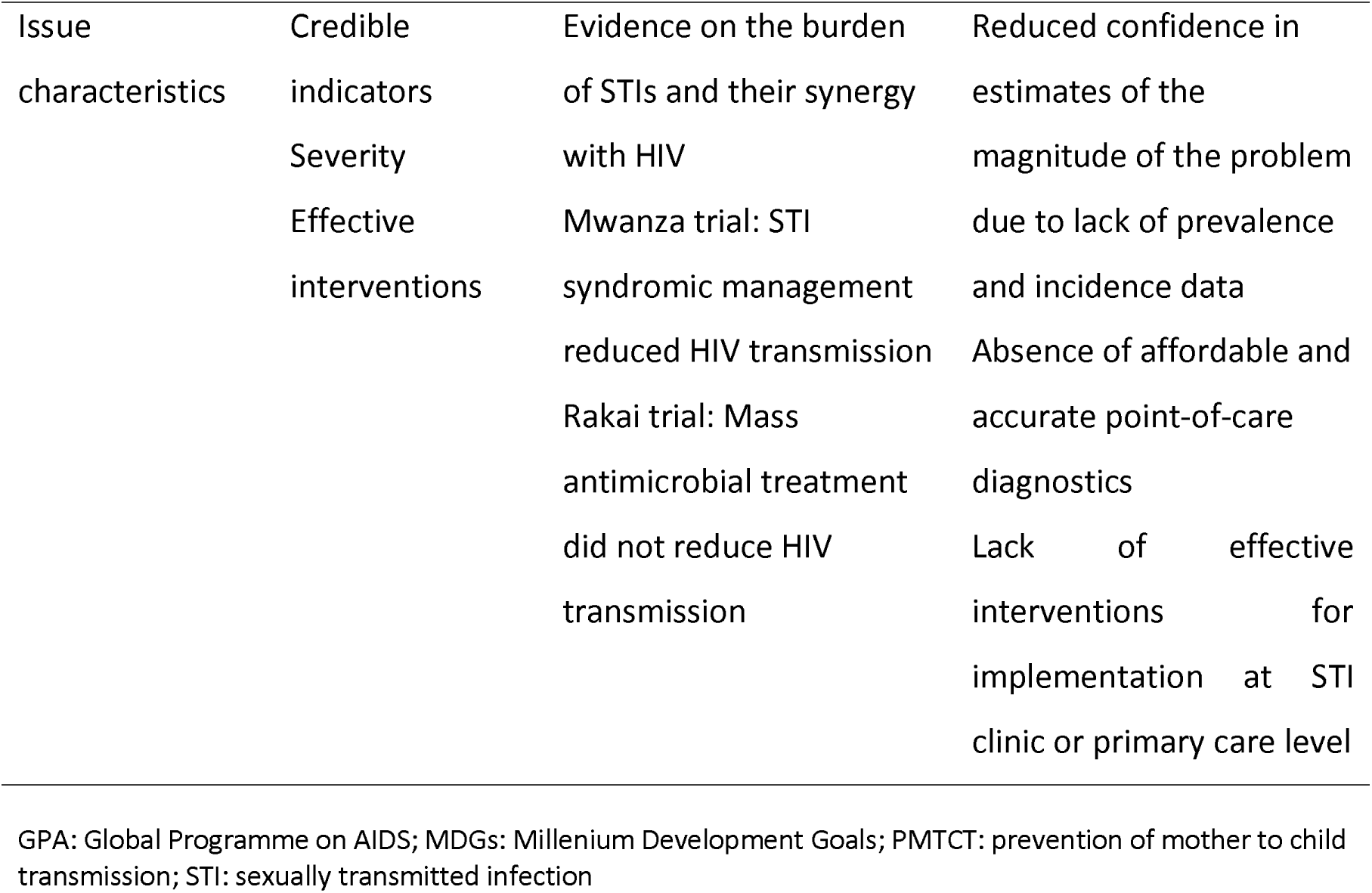
Factors affecting global political priority for controlling STIs.

## Discussion

Our study analyses the factors that have influenced the priority afforded to STIs by global health actors over time. STI control was high on the global health agenda during the late 1980s and 1990s, when the world was looking for cheap, effective and feasible solutions to the HIV/AIDS epidemic. At that time, a strong global policy community agreed on both the high burden of STIs and the potential of STI management to reduce the incidence of new HIV infections. However, as indicated through informant interviews, the level of priority decreased when research evidence did not find an impact of STI control interventions on HIV incidence. Since 2000, the global STI policy community has largely been characterised a loosely organised structure, absence of champions, undefined coordinating mechanisms, lack of compelling issue framings, and insufficient engagement of civil society. These factors, along with uncertainties surrounding the actual burden of STIs and cost-effectiveness of interventions, have contributed to challenges of achieving policy salience.

Our study suggests reasons for the gap between perceived and actual priority of STIs when examining the limited ODA allocation data. Most respondents believed that STI control had fallen off the global health agenda since the late 1990s and remarked on decreased levels of funding from major donors – responses did not appear to vary by gender identity or occupational history. This sentiment was often linked to the change in scientific evidence, with the findings of the Rakai trial^43,44^ and others,^47–49^ which broke the consensus that STI control interventions could reduce HIV transmission. Despite substantial estimates of the funds needed for broad STI control, there has been under-funding, compared to more targeted initiatives, like PMTCT of syphilis, HPV vaccination and treatments for antimicrobial-resistant gonorrhoea.^57^ The disconnect between perception and evidence could result from a constrained understanding of what constitutes STI control and where it is delivered. Informants talked generally about STI control without specifying infections or interventions. Those focused on curable STIs might overlook the priority given to HPV vaccination as it was widely promoted as cancer prevention. Sample bias might also have contributed to this finding, as three-fourths of our respondents first became engaged in STI control in the late 1980s and 1990s, likely reflecting the higher priority assigned to STIs during that period. Another limitation of our study is the underrepresentation of respondents from LMICs and the absence of participants from major donors.

Our study has identified factors to consider for those seeking to boost resources for STI control. Political science suggests that a “policy window” opens when three streams – policy, problem and politics – converge.^27^ There is first a need for the global STI policy community to recognise the importance of political decision-making as well as empirical evidence in driving policy attention and resource allocation. For instance, the PMTCT of syphilis programme’s alignment with MDGs 4, 5 and 6 strategically placed congenital syphilis control within a broader health and development narrative, capturing international policy attention and funding (the politics stream). This alignment, along with cost-effective interventions and concrete evidence of the disease’s global burden (the policy and problem streams, respectively), was driven by “political entrepreneurs” – Individuals from WHO, academia, and civil society. These stakeholders partnered to raise the salience of congenital syphilis, merging the three streams into a window of opportunity for increased priority.

Second, framing is crucial for policy prioritisation. Control of curable STIs was prioritised when framed as a means of achieving HIV control earlier in the epidemic. The evidence that STI management did not decrease HIV transmission dealt a blow to funding for non-HIV STIs. Subsequent STI control programmes that have achieved more financial and priority “success” have been framed as cancer control (HPV vaccination) and improving neonatal and maternal health (congenital syphilis). To elevate other STIs (e.g., chlamydia and gonorrhoea) on policy agendas, finding alternative framings beyond the “STI control” narrative is likely necessary.

Third, action is needed to address the STI community’s apparent lack of cohesion, advocates, champions, and politically strategic framings. The role of advocacy coalitions in global health is well described, particularly in the case of HIV and access to antiretroviral treatments. The field of STI control, however, appears to lack coordinated engagement with stakeholders beyond congenital syphilis and HPV vaccines. Efforts to identify and engage with a range of stakeholders across civil society, reproductive health advocates, adolescent health champions etc., will likely foster a strong and successful advocacy movement for STI control.

Fourth, the emergence of attention to UHC around 2015, along with an ongoing emphasis on health systems strengthening, may offer new opportunities for integrating STI control into broader health policy agenda. While UHC is essential for realising the right to health for all, limited resources necessitate priority setting to ensure fair and efficient resource allocation, especially for marginalised and vulnerable populations.^58^ Syndromic management and partner notification remain the main interventions available for controlling curable STIs in the general population in most countries,^59^ so efforts should be made to make sure that they are part of UHC.

Lastly, recognising that ODA contributes only a limited part of total STI financing, and considering the frequent exclusion of STI services from essential service packages, it is crucial to take measures at national level. These should include identifying reliable funding sources, establishing strategic coordination, and ensuring equitable service provision along with quality assurance.^15,37^

## Conclusion

Our study highlights the importance of recognising the political nuances in policy attention and resource allocation beyond empirical evidence, and understanding the roles that values, framing, coalitions and strategic management of evidence into processes can play. The rise of UHC since 2015 offers a promising avenue to integrate STI initiatives into broader health strategies, which will require a concerted effort to frame STI interventions appropriately (i.e., framing linked to a broader agenda beyond STIs), and forge connections with other communities and stakeholders focused on sexual and reproductive health agendas.

## Patient consent for publication

Not applicable.

## Data availability

There were no new datasets generated or analysed for this study. Due to the confidential nature of the interviews, transcripts are not accessible to the public. Supplementary material, including the questionnaire template, is in the online supplemental file.

## Acknowledgements

We extend our sincere gratitude to the informants who generously shared their insights and experiences, making this study possible.

