## Supplementary material for "Understanding the factors affecting global political priority for controlling sexually transmitted infections: a qualitative policy analysis": online supplemental file

### Supplemental file 1a: Information sheet

#### PARTICIPANT INFORMATION SHEET

**Interview with key stakeholder**

**Please read this information sheet before you decide to take part. Please ask if there is anything that is not clear.**

**Title of Study:**
Political prioritisation of the prevention and control of sexually transmitted infections: a global challenge

**Institute:**
Institute of Social and Preventive Medicine, University of Bern, Mittelstrasse 43, 3012 Bern, Switzerland

**Name and Contact Details of the Researcher:**

**Name and Contact Details of the Project Coordinator:**

1. **Invitation**
   You have been invited to take part in a research project about the political prioritisation of sexually transmitted infections (STIs) as a global health issue. Prior to deciding on whether to participate or not, it is important for you to understand why this research is being conducted and what participation will entail. Please take time to read the following information carefully and discuss it with others if you wish. Please ask the researcher if there is anything that is not clear or if you would like more information. Take time to decide whether you wish to take part.

2. **Who is organising and funding the research?**
   This research project is carried out by the Institute of Social and Preventive Medicine at the University of Bern and funded by the Swiss Network for International Studies.

3. **What is the project's purpose?**
   This research project focuses on agenda setting and policy formulation for the prevention and control of STIs as a global public health problem. The project aims to understand how, why and why not, STIs have been placed on the global policy agenda during different time periods. Through identifying the key features driving or hampering prioritisation of the issue, we seek to achieve a better understanding of the health policy process and provide recommendations for promoting STI control at both global and national levels. The study involves analyses of policy documents and media coverage, as well as in-depth interviews with key stakeholders.

4. **Why have I been chosen?**
   We have invited you to participate in the study based on your expertise and experience in health policy. We are interviewing around 25-35 people, identified from policy documents and other experts in the field; but none of the other experts are aware that we have invited you to participate.

5. **Do I have to take part?**
Your participation is completely voluntary. It is up to you to decide whether or not to take part in the study. If you do agree to take part, you will be asked to sign an **Informed Consent Form** to confirm that you understand the purpose of the study and what is expected from you. Meanwhile, you can withdraw at any time without giving a reason (as long as it is before the results have been published). If you decide to withdraw, you will be asked what you wish to happen to the data you have provided until that point.

6. **What will happen to me if I take part?**
If you choose to participate in the study, we will ask you to take part in a semi-structured interview during which questions about STI policy at both global and national levels will be asked. The interview will include questions such as "How is your organisation involved in the promotion of STI control?" or "Who played a major role in shaping agenda setting for STIs?" If there are questions you do not wish to answer, you can opt out of answering them. The interview will take about 30 to 60 minutes via phone or internet call at your convenience. Your confidentiality and privacy will be ensured.

7. **Will I be recorded and how will the recorded media be used?**
If permission is received from you, the interview will be audio-recorded. If you prefer to not record the interview, notes will be taken by the researcher. Audio-recordings will be transcribed by the research team, and the audio-records will be deleted 3 years after the project is completed. The data will be stored locally on password-protected computers and will only be accessible by the research team.

8. **Will my taking part in this project be kept confidential?**
All the information that we collect about you during the course of this project will be used for the study purpose only and kept strictly confidential. You will not be personally identifiable in any ensuing reports or publications. If we use any direct quotes from your interview, we will only identify your organisation (e.g. "WHO official" or "academic) stating no other personal features in order to avoid any possibility of you being identifiable in person. If we plan to use a (anonymised) quote from you, we will seek your permission before the findings are presented to an external audience. If you do not wish the quote to be used or information disclosed, we will not present or publish this information.

9. **What will happen to the results of the research project?**
The results of the study will be published as part of a project report and in peer-reviewed journals. The findings will also be presented and discussed in meetings and workshops with relevant stakeholders.

10. **What are the possible disadvantages and risks of participating?**
There are no direct benefits for you if you choose to participate in the study. There is a possibility of reputational risk if politically sensitive information is divulged. However, your participation will be kept unidentified to your institution and, as described earlier, you will be given an opportunity to see and approve any direct quotes used from your interview prior to presentation or dissemination of the study results. If you experience any discomfort or risk associated with your participation in the research project, please let the research team know.

11. **What are the possible benefits of taking part?**
There are no intended direct benefits for you from taking part in the study. However, it is hoped that the findings of the study will be relevant for promotors, policymakers, and other stakeholders working on the prevention and control of STIs.

12. **What if something goes wrong?**
If you have any questions arising from this Information Sheet or explanation already given to you, please ask the researcher before you decide whether to join in. Dr. Dadong Wu, or Tel: +86 (0)13823184820. You will be given a copy of this Information Sheet to keep and refer to at any time.

If you have questions, concerns, or complaints, or think this research has hurt you, please contact the responsible project coordinator: Professor Nicola Low, or Tel.: +41 (0)316313092.

**13. Ethical approval**

This study is not subject to ethical committee approval in Switzerland due to the reason that it does not fall under the Swiss Human Research Act, Art. 2, Paragraph 1. The clarification of jurisdiction is presented with this Information Sheet.

Your time and cooperation is highly appreciated!

### Supplemental file 1b: Consent form

**I confirm that I understand that by ticking or initialling each box below, I am consenting to this element of the study. I understand that it will be assumed that unticked or initialled boxes mean that I DO NOT consent to that part of the study. I understand that by not giving consent for any one element that I may be deemed ineligible for participation in the study.**

| **Study element** | **Tick Box** |
|---|---|
| I confirm that I have read and understood the **Participant Information Sheet** for the above study. I have had the opportunity to consider my participation and ask questions, which have been answered to my satisfaction. | |
| I consent to participate in the study as described and understand that I am free to withdraw at any time without giving a reason. I understand that if I decide to withdraw, any personal data I have provided up to that point will be deleted unless I agree otherwise. | |
| The study procedures have been explained to me and I understand them. I understand that all my personal information collected will be used for the study purpose only. | |
| I understand that all the data gathered in this study will be kept strictly confidential and that all efforts will be made to ensure that I cannot be identified. | |
| I understand the potential risks of participating. | |
| I understand the societal benefits of participating in research. | |
| I understand that the information I have submitted will be published as part of a project report and in peer-reviewed journals, but I will not be identified in person, only pseudo-anonymously through my organisation. | |
| a. I consent that written notes will be taken during my interview. **(Please choose between 8a or 8b)** | |
| b. I consent to that my interview will be audio-recorded and I understand that the recordings will be destroyed following transcription 3 years after study completion. **(Please choose between 8a or 8b)** | |
| I am aware of who I should contact if I wish to lodge a complaint. | |
| I understand that the study is not subject to ethical committee approval in Switzerland, and I have read the clarification of jurisdiction. | |

Name of participant    Date    Signature

____________________    ______________    ________________

Researcher    Date    Signature

____________________    ______________    ________________

#### Supplemental file 2: Project group members, “Political prioritisation of the prevention and control of sexually transmitted infections: a global challenge”

| Name | Affiliation |
|---|---|
| Nicola Low | Institute of Social and Preventive Medicine (ISPM), University of Bern |
| Hira Imeri | Institute of Social and Preventive Medicine (ISPM), University of Bern |
| Eva Cignacco | Bern University of Applied Sciences, School of Health Professions, Midwifery Division Bern Switzerland |
| Sarah Hawkes | Institute for Global Health, University College London |
| Dadong Wu | Affiliated Shenzhen Maternity & Child Healthcare Hospital, Southern Medical University, Shenzhen, China<br>Center for World Health Organization Studies, School of Health Management of Southern Medical University, Guangzhou, China |
| R Matthew Chico | Department of Disease Control, Faculty of Infectious & Tropical Diseases, London School of Hygiene and Tropical Medicine, London, United Kingdom |
| Kelvin Kapungu | Tropical Disease Research Centre, Ndola, Zambia |
| Mike Chaponda | Tropical Disease Research Centre, Ndola, Zambia |
| Mae Dirac | Institute of Health Metrics and Evaluation, University of Washington, WA, USA |
| Angela Kelly-Hanku | Papua New Guinea Institute of Medical Research, Papua New Guinea<br>The Kirby Institute, University of New South Wales, Sydney, Australia |
| Lisa Vallely | Papua New Guinea Institute of Medical Research, Papua New Guinea<br>The Kirby Institute, University of New South Wales, Sydney, Australia |
| Andrew Vallely | Papua New Guinea Institute of Medical Research, Papua New Guinea<br>The Kirby Institute, University of New South Wales, Sydney, Australia |
| William Pomat | Papua New Guinea Institute of Medical Research, Papua New Guinea |
| Melanie Taylor | US Centers for Disease Control and Prevention, Atlanta, Georgia, USA |
| Jane Rowley | Department of HIV, hepatitis and STIs, World Health Organization, Geneva, Switzerland |
| Nathalie Broutet | Department of Reproductive Health Research, World Health Organization, Geneva, Switzerland |
| Dianne Egli-Gany | Institute of Social and Preventive Medicine (ISPM), University of Bern |

### Supplemental file 3: Ethics waiver

**Kanton Bern**
**Canton de Berne**

Gesundheits-, Sozial- und Integrationsdirektion
Kantonale Ethikkommission für die Forschung

Murtenstrasse 31
3010 Bern
Bern
+41 31 633 70 70 (Telefon)
+41 31 633 70 71 (Telefax)

www.be.ch/gsi

Dorothy Pfiffner
+41 31 633 70 77


GSI-KEK, Murtenstrasse 31, 3010 Bern

Prof. Nicola Low
University of Bern, ISPM
Mittelstrasse 43
3012 Bern

**Clarification of jurisdiction**

**BASEC-Nr:** Req-2020-00269 **Date of receipt:** 10/03/2020

**Title:** Political prioritisation of the prevention and control of sexually transmitted infections: a global challenge

Please refer to the uploaded document for a description of the project.

**Result of clarification of jurisdiction**

☒ **Not responsible**: The project is not subject to ethical committee approval in Switzerland. Reason: The project does not fall under the Human Research Act, Art. 2, Paragraph 1.

☐ **Responsible:** Approval according to Human Research Act, Art. 2, Paragraph 1 is necessary in Switzerland. Please submit an application to the KEK according to www.swissethics.ch.

**Fee**: CHF 200.-- (Tariff code 6.0)

Date/Place: 23.03.2020/Bern

Prof. Dr. med. Christian Seiler
President

Dr. sc. nat. Dorothy Pfiffner
Head of the scientific secretariat

Req-2020-00269 1/1
